# BodyMirror Clinical MS™: A Multimodal Game-Based Digital Therapeutic for Remote Monitoring and Neurorehabilitation in Multiple Sclerosis

**DOI:** 10.64898/2026.03.06.26347719

**Authors:** Dr Zied Tayeb, Samaher Garbaya, Bernhard Specht

## Abstract

**Background:** Multiple sclerosis (MS) is a chronic neurodegenerative disease characterised by progressive neurological disability and heterogeneous symptom trajectories. Current clinical monitoring methods, including magnetic resonance imaging (MRI) and episodic neurological assessments, provide limited insight into subtle disease progression and real-world functional changes. Digital health technologies integrating multimodal biosignals and behavioural assessments may enable continuous monitoring and personalised rehabilitation for patients with MS.

**Objective:** This study aims to evaluate the clinical utility of the BodyMirror Clinical MS™ platform, a multimodal software-as-a-medical-device (SaMD) that combines wearable biosensors, neuroscience-based games, and machine learning algorithms to remotely monitor disease progression and deliver personalised neurorehabilitation for individuals with multiple sclerosis.

**Methods:** This study is a prospective, randomised, double-blind, controlled, multisite clinical trial enrolling 400 participants, including 300 individuals with multiple sclerosis and 100 healthy controls. MS participants will be randomly assigned (1:1) to either an adaptive neurorehabilitation intervention group or a control group receiving non-therapeutic digital activities matched for engagement and exposure. Participants will perform three 30-minute sessions per week over a 24-month period using the BodyMirror platform. The system integrates multiple biosignals, including electroencephalography (EEG), electromyography (EMG), inertial measurement unit (IMU) motion data, speech analysis, and behavioural performance metrics, to generate digital biomarkers of neurological function. The primary endpoint is change in Expanded Disability Status Scale (EDSS) score from baseline to 24 months. Secondary outcomes include changes in Multiple Sclerosis Functional Composite (MSFC), MRI brain volume, cognitive performance, patient-reported outcomes, adherence to digital rehabilitation, and health-economic outcomes.

**Conclusions:** This trial will provide the first large-scale clinical evaluation of a multimodal digital neurotechnology platform combining wearable biosensors and game-based neurorehabilitation for remote management of multiple sclerosis. If successful, BodyMirror Clinical MS™ may enable scalable remote monitoring, earlier detection of disease progression, and personalised digital rehabilitation for individuals living with MS.

## 1. Protocol Synopsis

**Study Title:** BodyMirror Clinical MS™ Remote Monitoring and Rehabilitation Trial (R-MMS Trial)

**Primary Objective:** To evaluate whether the BodyMirror Clinical MS™ platform improves neurological outcomes and provides clinically valid digital biomarkers for monitoring disease progression in patients with multiple sclerosis.

**Secondary Objective:** To assess the efficacy of the BodyMirror game-based neurorehabilitation programme in improving cognitive, motor, and quality-of-life outcomes.

**Design:** Randomised, double-blind, controlled, parallel-group trial (24-month duration).

**Sample Size:** 400 participants (300 MS patients:150 per group, 100 healthy controls).

**Registration:** This clinical trial is registered on **ClinicalTrials.gov** prior to participant enrollment.

**MS Population:** Adults aged 18-65 with confirmed RRMS.

**Primary Endpoint:** The primary endpoint is the change in Expanded Disability Status Scale (EDSS) score from baseline to Month 24 between the intervention and control groups.

### Key Secondary Endpoints

- Change in **Multiple Sclerosis Functional Composite (MSFC)**
- Change in **brain volume measured by MRI**
- Change in **cognitive performance scores**
- Change in **patient-reported quality of life**
- Adherence to a digital rehabilitation programme

We aim to conduct an innovative multi-site clinical trial to demonstrate further the effectiveness of a new software-as-a-medical device (SaMD), so-called BodyMirror Clinical MS™, whose purpose is the remote monitoring of silent MS progression and patient rehabilitation. Our objectives:

**Primary Objective 1:** Test if the SaMD innovation can indeed monitor a) silent disease progression and b) monitor symptoms and patient response to treatment. Results from the monitoring group will be benchmarked in terms of the relative diagnostic accuracy of the innovation in monitoring a) disease progression, b) patient response to treatment (how well it can perform in comparison to the gold standard). We will evaluate the correlation between BodyMirror Clinical MS™ metrics and standard clinical measures (MRIs, standard clinical tests, etc.). Early markers of disease progression will be identified. This endpoint will allow us to clinically validate the proposed SaMD and its effectiveness by demonstrating its accuracy, sensitivity, reliability, and robustness to remotely monitor MS progression and treatment response across different patients recruited in the proposed multi-site clinical trial. The focus will be on silent disease progression and treatment response. Our neuromorphic algorithms and games will extract and recognise patterns from each digital biomarker while playing the different games. We will analyse the difference (delta & changes) before and after a specific medication/treatment, as well as regularly monitor symptoms and quantify the disease trajectory. The performance, robustness, and reliability of MyelinZ’s SaMD and the accuracy of its software results will be compared to MS gold standard tests (MRI scans, classical clinical tests such as SDMT, etc.) and the EDSS score.

**Primary Objective 2:** Test if the SaMD could enhance cognitive abilities: cognitive & Physical rehabilitation using group 2. This will be performed by comparing the change in EDSS scores over 24 months between intervention and control groups. We will also assess changes in MSFC scores, brain MRI volume, quality of life, and other cognitive functions.

We will compare changes in:

- Multiple Sclerosis Functional Composite (MSFC) scores
- Brain volume on MRI
- Quality of life measures
- Cognitive function
- Patient adherence
- Patient satisfaction

By dividing patients into two groups: one for passive monitoring and another for participating in our neurorehabilitation games, our goal is to validate these rehabilitation games, which aim to enhance working and short-term memory, visual processing speed, attention, speech fluency and articulation, upper and lower-limb dexterity, mood, etc. The games incorporate neurofeedback and biofeedback to promote neuroplasticity. MRIs, performance in classical tests, and the patient’s self-reported outcome and impact on their daily life will be used as objective measures to validate the rehabilitation programme and concept.

**Secondary Objective 1:** Collect health economics data to demonstrate cost benefits + To assess the efficacy of the BodyMirror game-based neurorehabilitation programme in improving cognitive, motor, and quality-of-life outcomes.

### Overall Study Design

- Prospective, randomised, double-blind, parallel-group, controlled trial
- Blinding: Participants, outcome assessors, and data analysts will remain blinded to treatment allocation.
- Randomisation: 1:1 ratio

**Sample Size**: 400 participants (300 MS patients: 150 per arm) and 100 healthy participants

### Study Duration

- Enrollment Period: 6 months
- Treatment & Monitoring Period: 24 months
- Total Study Duration: 30 months

**Study Population**: Adults with confirmed relapsing-remitting multiple sclerosis (RRMS) (40%) and secondary progressive multiple sclerosis (60%). 100 healthy participants.

**Primary Endpoint**: The primary endpoint is the **change in Expanded Disability Status Scale (EDSS) score from baseline to Month 24** between the intervention and control groups.

### Secondary Endpoints

- Change in **Multiple Sclerosis Functional Composite (MSFC)**
- Change in **brain volume measured by MRI**
- Change in **cognitive performance scores**
- Change in **patient-reported quality of life**
- **Adherence to a digital rehabilitation programme**

## 2. About The Sponsor

MyelinZ has developed a non-invasive brain-machine interface platform that integrates artificial intelligence (AI) and neurogames to monitor and improve brain health remotely. We are sponsoring an international, multi-site clinical trial across Europe, the US, and the UK to validate a novel game-based neurotechnology for the remote monitoring and rehabilitation of patients with neurodegenerative diseases, specifically multiple sclerosis. Our core novelty is the gamification of neuroscience and neurology. Patients play a set of games to assess their brain and body health while their brain activity is captured. Based on AI analysis, we create a game-based treatment plan aimed at alleviating symptoms and enhancing the cognitive and physical abilities of patients. The company has five filed patents in the US.

## 3. Background and Rationale

**Disease Background:** MS is a chronic autoimmune disease characterised by demyelination and neurodegeneration in the central nervous system.

More than 2.8 million people have MS, and the disease causes a reduction in life expectancy of 7–14 years [1]. Unlike any other brain disorder, MS is commonly diagnosed in the 20s & 30s and is the most common cause of disability in younger adults [2]. Recent statistics have shown that more than 42% receive inadequate treatment within the first stage of the disease, known as RRMS. Moreover, over 50% of MS patients suffer from silent disability progression [3]. Although MS is incurable [4], adequate treatment can manage symptoms, treat relapses, and slow the progression of RRMS. With more than 25 possible medications for RRMS (approved by the FDA) [5] and a no-one-size fits all approach, it has become imperative to remotely monitor medication effectiveness and, more importantly, monitor silent disease progression and any cognitive decline in RRMS patients [6]. To date, MRI is the only available method to monitor the progression of MS, but it has several limits (costs, feasibility, patients need to be hospitalised, less capable of detecting atrophy, etc.) [7]. Further, MRI, although often used as a non-clinical endpoint in clinical trials, cannot predict treatment effectiveness or predict relapses. Similarly, other clinical routines and tests to monitor progression, such as the EDSS clinical score, are not objective and inaccurate as they usually only involve visual inspection of gait and walking ability [8].

**Current Challenges in MS Management:** Limited monitoring tools, difficulty tracking disease progression, need for engaging rehabilitation methods.

- Limited access to regular monitoring
- Lack of engagement in rehabilitation programmes
- Difficulty in tracking subtle disease progression
- Need for frequent clinical visits
- Limited objective measurement tools

Hence, regular monitoring and rehabilitation are crucial for managing disease progression and maintaining functional abilities.

**Proposed Approach:** Inspired by his mother’s battle with multiple sclerosis and her disability, Dr Tayeb pursued a PhD in neuroscience and co-founded MyelinZ to pioneer personalised remote monitoring and cognitive rehabilitation for MS patients. MyelinZ has developed a novel software-as-medical-device (SaMD) for remote (home-based) monitoring of neurological disorders, including MS patients. The SaMD is a software platform (non-invasive, hybrid, and closed-loop brain-computer interface (BCI)) that combines mobile cognitive (neuroscience-based) games, a wearable brain sensor (wireless, gel-free, clinically certified), a biologically inspired machine learning engine for real-time (on-device) data processing, and a clinical dashboard for clinicians. While patients play MyelinZ’s cognitive games on their phones for a few minutes, different biosignals are captured. The device captures a myriad of biosignals, namely speech (vocal patterns), EEG (cognitive and central nervous system digital biomarkers), mood and emotions using facial expression & voice, EMG (peripheral nervous system digital biomarkers), gait and body movements (using Inertial measurement unit), skin temperature, heart rate through PPG, as well as cognitive performance (using game performance metadata). We gamify the neurological assessment process to ensure players’ compliance and willingness to perform such sessions at home. Collected biosignals are processed in real-time (on-device) using MyelinZ’s proprietary machine-learning and translated into digital biomarkers & a novel clinical score, shared via an online (PDF) medical report along with clinical recommendations. Such digital biomarkers can help health professionals detect early and rapid disease worsening, which allows them to prescribe adequate treatment, as well as provide a comprehensive overview of the patient’s health status anytime, anywhere, thereby improving monitoring and prediction of the impact of disease on people’s lives, including disability trajectories. Based on the monitoring scores, a personalised game-based neurorehabilitation programme is created. These cognitive rehabilitation games target specific symptoms, such as cognitive decline, motor impairment, and mood disturbances. By leveraging neuroplasticity, the neurogames will complement pharmacological treatments. Such SaMD will chart a route ahead for the next generation of brain disorder remote monitoring platforms.

**Study Rationale:** Assessing BodyMirror’s potential for improving outcomes through game-based monitoring and rehabilitation, focusing on its unique multimodal approach and remote monitoring capabilities. The BodyMirror system integrates multimodal biosignal acquisition, digital behavioural assessment, and adaptive neurorehabilitation through game-based tasks. By combining EEG, EMG, movement sensors, speech analysis, and behavioural metrics, the platform generates digital biomarkers that may provide earlier detection of neurological changes compared to traditional episodic clinical assessments.

### I. Rationale for Including Healthy Controls

A. Scientific Value

1. Establish normative data for BodyMirror metrics
2. Validate game-based assessments
3. Differentiate disease-specific from normal variation
4. Control for learning effects
B. Clinical Relevance

1. Define performance boundaries
2. Establish clinically meaningful changes
3. Validate sensitivity and specificity
4. Support diagnostic potential

### II. Rationale for Double-blind Trial

Double-Blind Trial:

- Both participants and researchers are unaware of which group the participants are in.
- A third-party administrator or an automated system typically manages the assignment and monitoring.
- This design reduces both participant and researcher bias, leading to more reliable results.

## 4. Previous Work

### Previous Clinical Research Work & Novelty

In recent times, we have unequivocally witnessed a push towards digitising the healthcare system. Topics such as RPM, digital health, and their use to monitor neurological disease progression have gained momentum and popularity. Notwithstanding the considerable advances that have been made in adopting such technologies and using them in the context of mental health or even a few neurodegenerative disease monitoring, they have not been widely used in the context of remote management and treatment of MS. In the same vein, given that MS is a very individualised disease to manage, there are numerous challenges, yet opportunities associated with using digital health technologies for remote MS monitoring. Here, we review some of the previous work and clinical attempts performed over the last decade (mainly and predominantly, hospital-based monitoring) en route to using digital health for MS monitoring and management. Throughout this extensive review, we shine a light on various monitoring methods that hold the potential to be measured in a home environment, including EEG and evoked potentials (e.g., MEP, SSEP, VEP, EMG, IMU, and speech analysis. However, we stress the fact that, to the best of our knowledge, combining such digital biomarkers altogether for MS monitoring (progression, treatment response, or both) has never been performed before. Along the same lines, almost all the studies carried out before and mentioned in this section are hospital-based, and there have not been any (home-based) digital devices for remote MS monitoring.

- **EEG-based clinical studies:** The detection of VEP from EEG signals was used in 59 patients, and the quantification of its amplitude and latencies between different MS patients was studied in 40 patients **[33]**. In another study **[34]**, authors showed that clemastine fumarate treatment reduced VEP latency by 1.7ms, albeit the study was only conducted with 25 patients. In the same vein, in another study **[35]**, higher doses and frequency helped to reduce the relapse rate and, subsequently, the VEP latency. At the same time, there were no significant changes in radiological and electrophysiological findings **[35]**. Similarly, a different clinical study **[36]** showed that the VEP latency (known as P100) is significantly higher in MS patients, and secondary progressive MS patients had the highest delay. Similarly, authors in **[37]** found that latencies of VEP in EEG signals are significantly altered in MS and showed that changes in VEPs over time have been observed. The changes may contain valuable information, but within the paper, there was no correlation analysis (e.g., medication). In a clinical study published in 2021 **[38]**, the authors showed that prolonged VEP latency was significantly associated with poorer performance in multiple cognitive domains (poorer memory, executive function, attention, visual-spatial processing, information processing speed, motor skills, and global cognitive score) in MS patients but no investigation of the correlation with disease progression or treatment response was performed.
- **EMG, gait, and IMU-based clinical studies:** In this study **[39]**, when analysing EMG data, MS patients have a lower relative decrease in torque during repeated maximal concentric contractions than healthy subjects, reflecting lower fatigability. In another study **[40]**, within a short follow-up interval, a multi-sensor algorithm combining EMG and IMU distinguished progressive from relapsing MS and captured changes in limb function. Authors in **[41]** showed by analysing EMG and IMU data that smoothness and grip force control are most affected in pwMS. Those abnormalities impact the time to complete the goal-directed task. Anomalies in movement patterns and grip force control were consistently found even in pwMS with clinically average gross dexterity and grip strength. Along the same lines, another study **[42]** on EMG and IMU data revealed significant improvements after intensive neurorehabilitation in most of the clinical assessments, cadence, and velocity of the instrumental gait analysis, paralleled by amelioration of thigh co-activation on the less-affected side. Authors highlighted that improved gait stability and balance could be the result of reduced inappropriate muscular co-activation of the lower limbs, indicating the importance of integrating kinematic and muscle activity measurements to evaluate and interpret treatment effects objectively.
- **Speech-based clinical studies:** In this study **[43]**, the authors revealed that speech biomarkers from reading a text could be used to extract features and predict the health status (MS or Healthy) with 82% accuracy using a simple machine learning model (random forest), although no monitoring was performed. In a different study **[44]**, the authors showed that patients with PMS had reduced articulation and speech rates than controls and patients with RRMS. Similarly, another study **[45]** showed that articulation rate in speech is correlated with bilateral white and grey matter loss, as well as brain atrophy and EDSS. In **[46]**, the authors showed that the DSI and Centralisation Ratio significantly differed between controls and MS patients. Furthermore, the formant Centralisation Ratio correlates with EDSS and disease duration, and the authors pinpointed that the articulation subsystem changes might be suitable for monitoring the progression of the disease. Along the same lines, the authors of **[47]** found that expiratory times in speech were significantly correlated with the EDSS scores and could be used as a measure of severity and progression in MS. Overall, the study revealed that MS patients have lower maximum expiratory times and maximum phonation times compared to the Control Group.

## 5. BodyMirror System Specifications

**System Overview:** BodyMirror is a multi-modal, AI-driven system comprising:

- **Game Suite:** 16 neurogames designed for cognitive, physical, and balance assessment and rehabilitation.
- **Sensors:** EEG for brain activity, EMG for muscle monitoring, and IMU for movement data (band). The EEG Headband has already passed all safety requirements (a four-channel EEG, non-invasive, gel-free, Bluetooth communication, Sampling rate: 256 Hz, Resolution: 24-bit). The EMG band has already passed all safety requirements (Sampling rate: 1000 Hz, one bipolar EMG channel). **Sensors’ specifications are provided in the appendix.**
- **AI-driven progress tracking algorithms:** algorithms for data processing.
- **MyelinFace:** A patient app where they can play the prescribed games.
- **MyelinBoard:** A medical dashboard where results and interpretations can be accessed and where games can be prescribed.
- A tablet will be provided to play the games, although the games can be played on any mobile device (IOS or Android).
- All the games, instructions, results, MyelinFace, and MyelinBoard are available **in five languages: English, French, German, Italian, and Spanish.**

### Process

The BodyMirror SaMD combines twenty mobile cognitive games, a wearable (visor-like) sensor to capture various bio signals, a biologically-inspired machine learning engine to interpret the data in real-time, and a clinical dashboard. You start by playing our neuroscience-based games for a few minutes on your phone, at home. The games include one to capture vocal patterns, one to measure muscle fatigue, one to measure your gait and body movements, measure visual pathways in the brain using neuroscientific patterns, memory, attention decline, motor responses, wordle-like games to monitor and improve hand dexterity, a racing game, and endless runner games to measure foot tapping and lower-limb impairments, etc. Behind each game, there is a solid neuroscientific clinical protocol which has been validated. You first wear a visor-like sensor to capture brain activity and a belt that captures your muscle and body movement. Our core novelty is the gamification of neuroscience and neurology. You then play a set of games to assess your brain and body health, we capture your brain activity through EEG, muscle activity through EMG, gait, mental health, speech, etc.). We combine performance-based data from the games with sensor-based data from the body. Thereafter, we establish your baseline (machine learning algorithms will process all the data to develop an overall assessment of your current brain health status) and prescribe a game-based treatment plan whereby you play our designed games to help alleviate symptoms and enhance patient’s cognitive health (memory, visual processing speed, attention, gait, muscle strength, mood, speech, etc.) using neuroplasticity. We leverage AI to automate the process of prescription (the number of games, frequency, intensity, duration of the monitoring and rehab programmes, etc.). The rehab approach will complement pharmacological treatments. All the patterns are automatically extracted, quantified, and weighted to form a reliable monitoring score. Thereafter, the clinical scores alongside the detected digital biomarkers are used to generate real-time clinical recommendations that will be shared with health professionals.

## 6. Trial Design

### A. Baseline Establishment

- All patients will undergo a four-week baseline establishment process, whereby they will play all games **(three times a week) for four weeks**. Sessions will be divided into cognitive, balance & coordination, and physical assessments and level progress will be incremental (every game has 10 levels with an increasing complexity between levels). After four weeks, the patient’s performance will be assessed and main symptoms (deficits) will be identified, and a personalised rehabilitation programme will be created. The personalisation will determine the intensity and frequency of playing the games. During the baseline period all games will operate in **non-adaptive mode** to maintain allocation concealment.

### B. Classical Clinical Tests

This is a list of all the classical clinical tests that a patient needs to undergo at the start of the trial and every quarter thereafter. All those tests have been computerised and are available on the MyelinBoard:

**Physical Tests:**

- Multiple Sclerosis Functional Composite (MSFC)
- Timed Up and Go (TUG)
- Box and Block Test (BBT)
- Berg Balance Scale
- Timed 25-Foot Walk (T25FW)
- 9 Hole Peg Test (9HPT)
- Paced Auditory Serial Addition Test (PASAT)
- Symbol Digit Modalities Test (SDMT)

**Questionnaires/Scales:**

- Fatigue Assessment Scales
- Modified Fatigue Impact Scale
- Patient Health Questionnaire-8 (PHQ-8)
- Voice Handicap Index (VHI)
- Generalized Anxiety Disorder-7 (GAD-7)
- Multiple Sclerosis Impact Scale (MSIS-29)
- Tremor and Coordination Scale
- Bain Score for Tremor Severity

**Neurological Tests:**

- Expanded Disability Status Scale (EDSS)
- Relapse assessment

For imaging and laboratory tests, they will be conducted at the start of the trial every year thereafter.

**Imaging & Laboratory Tests:**

- Magnetic Resonance Imaging (MRI)
- Spinal Magnetic Resonance Imaging (MRI)
- Kappa Free Light Chains (KFLC)
- MRZ Reaction Data
- Neurofilament Light Chains (NFLs)

### C. Study Type

#### Overview

- Design: Prospective, randomised, double-blind, parallel-group trial
- Duration: 24 months
- Blinding: Evaluating physicians blinded to treatment allocation
- Randomisation: 1:1 ratio using computerised block randomisation

#### Trial Arms

**Intervention Group (n=150)**

**Participants receive:**

- Adaptive neurorehabilitation games
- Difficulty adjusted by AI
- Personalised training protocol

**Control Group (n=150)**

Participants receive:

- **Non-therapeutic digital activities** matched for:

- session frequency
- game duration
- visual design
- user interaction

These games:

- **contain** no targeted neurorehabilitation algorithms
- do not adapt the difficulty based on performance

**Blinding Procedures:** This study will implement a **double-blind design**.

Participants will be randomly assigned to either the therapeutic neurorehabilitation programme or a control digital activity programme that is visually and interactively identical but lacks adaptive therapeutic algorithms. Participants will not be informed of the functional differences between programmes. Investigating clinicians responsible for outcome assessments will remain **blinded to treatment allocation** throughout the study. Data analysts will receive **coded datasets without group identifiers** until the statistical analysis is completed.

**Key benefit**

Both groups:

- play games
- same schedule
- same interface

**Schedule Overview:** Screening (baseline establishment) period (−4 to 0 weeks), Randomisation (week 0), Game-based Treatment/Monitoring (weeks 1-96), Clinical Assessments every 3 months (using the list of classical clinical tests below), MRI at baseline, 12, and 24 months.

- **Blinding:** Participants, outcome assessors, and data analysts will remain blinded to treatment allocation.

**Study Schedule:**

- Week -4 to 0: Screening & Baseline Establishment
- Week 0: Randomisation
- Weeks 1-96: Game-based Treatment/Monitoring
- Months 3, 6, 9, 12, 15, 18, 21, 24: Clinical Assessments
- Months 0, 12, 24: MRI Scans (and laboratory tests)

### D. Study Population

Total N = 400 participants

1. MS Participants (n=300)

├── Rehabilitation Group (n=150)

└── Secondary progressive MS (n=90)

└── RRMS (n=60)

└── Monitoring Group (n=150)

└── Secondary progressive MS (n=90)

└── Early SPMS (n=45)

└── Established SPMS (n=45)

└── RRMS (n=60)

└── Early RRMS (n=45)

└── Established RRMS (n=45)

2. Healthy Controls (n=100)

├── Age-matched (n=50)

Young Adults (n=50)

**Rationale for 60% SPMS / 40% RRMS Distribution**

**Clinical Relevance**

- SPMS represents more advanced disease state
- Higher need for monitoring progression
- Greater potential benefit from rehabilitation
- More challenging to treat conventionally

**Scientific Considerations**

**Advantages of Higher SPMS Proportion:**

- Better assessment of progression monitoring
- More challenging test of rehabilitation efficacy
- Higher potential for detecting subtle changes
- Greater need for alternative interventions

**Statistical Power**

**Benefits:**

- Larger SPMS cohort (n=180 total)
- Better powered for progression detection
- More robust assessment of intervention effect
- Stronger subgroup analyses capability

**Monitoring Objective**

SPMS Focus (n=90 per arm):

- Better detection of subtle progression
- More frequent progression events
- Higher likelihood of detecting changes
- More challenging test of monitoring system

RRMS Component (n=60 per arm):

- Earlier disease stage monitoring
- Different progression patterns
- Relapse detection capability

**Rehabilitation Objective**

SPMS Focus:

- Greater rehabilitation need
- More challenging intervention test
- Higher potential impact
- More diverse symptom profile

RRMS Component:

- Early intervention potential
- Prevention focus
- Different rehabilitation targets

### E. Matching Criteria

1. Primary Matching Factors:

- Age (±3 years)
- Gender
- Education level
- Computer literacy
2. Secondary Matching:

- Handedness
- Geographic location
- Socioeconomic status

### F. Inclusion & Exclusion Criteria

**Inclusion Criteria (Patients):**

1. Age 18-65 years
2. Confirmed RRMS diagnosis (McDonald criteria 2017)
3. EDSS score 0-6.5
4. Disease duration ≥1 year
5. Stable medication regimen for ≥3 months
6. Internet access and device compatibility
7. Ability to provide informed consent
8. ● SSPM are patients whose EDSS score is between 6 and 7 and whose medical examinations have shown worsening symptoms and progression (change of at least 1 point of EDSS in the last 12 months).
9. ● RRMS patients will be selected according to McDonald diagnostic criteria. Medical history and previous clinical data assessment (EDSS, MRI scans, etc.) will be used for the selection of these patients.

**Exclusion Criteria (Patients):**

1. Other neurological conditions
2. Recent relapse (<3 months)
3. Planned medication changes
4. Severe cognitive impairment
5. Severe visual/motor impairment
6. Participation in other clinical trials
7. Pregnancy or planned pregnancy

Inclusion Criteria for Healthy Controls:

1. General Health:

- No neurological conditions
- No psychiatric conditions
- No chronic medical conditions
2. Specific Requirements:

- Normal or corrected-to-normal vision
- No motor impairments
- No cognitive impairments
- No history of substance abuse

**Exclusion Criteria**

1. Medical Conditions:

- Any neurological disorder
- Severe psychiatric conditions
- Uncontrolled medical conditions
2. Other Factors:

- Regular medication affecting CNS
- History of head trauma
- Substance use disorders

## 7. Assessment Schedule

### A. Healthy Controls Protocol

Baseline Assessment:

├── Clinical Measures

│ ├── Standard neurological examination

│ ├── Cognitive testing

│ └── Quality of life measures

├── BodyMirror Assessment

├── A first supervised BodyMirror training session (1 hour)

│ ├── All 22 games for four weeks

│ ├── Sensor data collection

│ └── Performance metrics

└── Follow-up Schedule

├── Fortnightly: Remote monitoring

├── Quarterly: Clinical assessment

**└──** Annual: Comprehensive evaluation

### B. Patients Protocol

Baseline Assessment:

├── MRI Scans, Blood Tests, and Lab tests

├── Clinical Measures (reported above)

│ ├── Standard neurological examination

│ ├── Cognitive testing

│ └── Quality of life measures

├── BodyMirror Assessment

├──A first supervised BodyMirror training session (1 hour)

│ ├── All 22 games for four weeks

│ ├── Sensor data collection

│ └── Performance metrics

└── Follow-up Schedule

├── Forntightly Remote Monitoring Sessions for group 1 (passive)

├── Weekly rehabilitation programme with increasing complexity and intensity and monitoring Sessions for group 2

├── Quarterly: Clinical assessment

**└──** Annual MRI Scans, Blood Tests, and Lab tests

**└──** Annual: Comprehensive evaluation and satisfaction surveys

**└──**Annual cost-effectiveness analysis

Both groups:

- 3 sessions per week
- 30 minutes per session
- same duration

---> Only difference: algorithm inside the games

**Intervention Schedule:** Participants in both arms will complete **three 30-minute sessions per week** using the BodyMirror platform over the 24-month study period. The intervention group will receive **adaptive neurorehabilitation games designed to stimulate cognitive and motor functions through progressive difficulty adjustments**. The control group will receive **non-adaptive digital activities designed to control for engagement, screen exposure, and motor interaction without targeted therapeutic mechanisms**.

**Healthy Control Group:**

1. Fortnightly sessions:

○ 3 fixed passive & remote monitoring sessions every two weeks to go all through the monitoring games

○ 30 minutes (max) per session

○ No Increase in levels or complexity compared to the baseline

○ Progress monitoring

○ The same monitoring games

### C. Comparative Assessments

1. Cross-sectional Analysis:

○ Game performance metrics

○ Cognitive measures

○ Physical assessments

1. Longitudinal Analysis:

○ Learning curves

○ Performance stability

○ Age-related changes

## 8. Study Procedures

### 8.1 Screening Procedures

1. Medical history review
2. Physical examination
3. Neurological examination
4. EDSS assessment
5. Cognitive testing
6. MRI scan
7. Laboratory tests

### 8.2 Randomisation

- Computerised block randomization
- Stratification by:

- Age
- EDSS score
- Disease duration

### 8.3 Blinding Procedures

Multi-level Blinding:

1. Patient Level

├── Neutral game interfaces

├── Standardised instructions

├── Identical app versions

└── Generic feedback

2. Investigator Level

├── Blinded assessment teams

├── Separate game-based treatment teams

├── Independent evaluators

└── Standardised protocols

3. Data Analysis Level

├── Coded group assignment

├── Blinded interim analysis

├── Independent statisticians

└── Sealed allocations

### 8.4 Objective Measurements

Data Collection:

1. Automated Metrics

├── Game performance data

├── Sensor measurements

├── Correlation between game performance data and sensor measurements

├── Response times

└── Error rates

2. Standardised Assessments

├── Validated instruments

├── Digital recordings

├── Multiple raters

└── Central review

## 8. Safety Monitoring

1. Adverse event reporting
2. Monitored Events: Adverse effects, MS progression, and new symptoms.
3. Monitor game-related complications (very unlikely or non-existent)
4. Regular safety assessments
5. Protocol deviation monitoring
6. **Independent Review Committee:** Quarterly reviews, with predefined stopping rules for risk.
7. Real-time Monitoring

a. Automated alerts
b. Safety thresholds
c. Emergency procedures

## 9. Data Management

### 9.1 Data Collection

1. Electronic Case Report Forms (eCRFs)
2. BodyMirror system data
3. MRI data
4. Clinical assessment data
5. Patient-reported outcomes

**Data Sources:** BodyMirror system logs, eCRFs, MRI, and patient-reported outcomes.

**Security:** Encryption, backup protocols, and access control to ensure data integrity.

### 9.2 Data Quality

1. Data validation procedures
2. Query management
3. Quality control checks
4. Audit trail

**Validation:** Checks for data accuracy, quality control, and monitoring logs.

### 9.3 Data Security

1. Encryption protocols
2. Access controls
3. Backup procedures
4. Privacy protection
5. Additional Measures:

- Technology familiarity questionnaire
- Gaming experience assessment (**already available on the app**)

### 9.4. Quality Control Updates

1. Standardisation:
2. Equipment calibration
3. Testing environment
4. Administration procedures
5. Data collection methods
6. Validation Steps:

- Cross-verification of health status
- Confirmation of eligibility
- Regular health monitoring
- Documentation requirements

### 9.5 Monitoring

1. Site initiation
2. Regular monitoring visits
3. Remote monitoring
4. Close-out visits

### 9.6 Training

1. Investigator training
2. System training
3. Assessment standardisation
4. GCP compliance

### 9.7 Data Problems Handling

- Missing Data: We will establish various procedures to minimize and handle missing data, such as implementing strategies for data collection and quality control, utilizing appropriate statistical techniques for imputation or managing missing data in the analysis, and documenting the approach taken.
- Unused Data: Unused data refers to data that was collected but not included in the final analysis. The reasons for unused data should be documented, ensuring transparency and reproducibility of the study.
- False Data: We will implement specific procedures to detect and address potential inaccurate data, including data validation checks, data quality control measures, and investigation of any suspected anomalies or inconsistencies in the data. Software algorithms will be developed and used to scrutinise data quality.
- **Findability:** data/research outputs from the project will be stored and appropriately catalogued in trusted OpenAire- and Zenodo-compliant repositories, and to relevant European data spaces (e.g., IDS), AI marketplaces (e.g., AI Assets Catalog of the AIOD platform), and Cloud infrastructures (e.g., European Open Science Cloud). Links to these repositories will be provided on the project’s web page. The outputs will be assigned persistent and unique identifiers and metadata to maximise their findability and identification through search engines.
- **Accessibility:** The use of these repositories provides the ability to keep the data available afterward, adhering to OpenAire guidelines in Horizon Europe, and, thus, to the EU Commission, Open access & Data management guidelines, following the principle “as open as possible, as close as necessary”. The used repositories will be included in registries of scientific repositories, such as DataCite and Databib, to increase the accessibility of the obtained results. Likewise, R-MMS will support the publication of data in open data portals, i.e., portals that operate at national, regional or municipal level (e.g., http://open-data.okfn.gr), and EU data portals (e.g., https://open-data.europa.eu/en/data). In case of data not being public, Article 19 and Article 22 of Commission Decision 2015/444 on the security rules for protecting EU classified information (March 2015) will be followed, being considered in the DMP. Although R-MMS will work towards open access, specific project results could also be protected and exploited, and, thus, IPR handling rules will be given (Section 2.2.5).
- **Interoperability:** existing standards and formats to specify, describe, and classify datasets and data types, and their associated metadata, will be adopted to ease interoperability, reusability, searching, filtering, and analysis activities. Metadata will be open (under CC0 or similar) and developed to meet the Metadata Quality Assurance of https://data.europa.eu.
- **Reusability:** the adopted strategy towards open access sharing in trusted repositories, EU data spaces (IDS) and marketplaces (Assets Catalog of AIOD…), along with a licensing strategy that fosters data sharing and re-usability, will make the project outputs (data, software…) highly reusable. Furthermore, the AutoAI system will strongly contribute to democratise the use of the released software, fostering the adoption and re-use of R-MMS solutions (Table 1). Curation and storage/preservation costs: all issues related to data management will be thoroughly detailed in the Data Management Plan (DMP). Data curation and recommendations will be defined by the consortium. Data will be collected, produced, and preserved (by covering associated costs, if any) in line with GDPR. On top of this, it is planned to build the outcome of the project partially on existing open-source tools. Therefore, we will also publish our tools and systems as an open-source project, preferably on GitHub. We will do that when a component is sufficiently mature, not later than the end of the project. To ensure reproducibility of the results, R-MMS consortium will adhere to agreed standards of research integrity, including transparent research design, the robustness of statistical analyses, and addressing negative results.

### 9.8 Data Management Plan

During the first three months of the trial, a Data Management Plan (DMP) will also be put in place. An “intranet” platform will be set up to ensure the storage of the data collected during the project and the backup of them. Each work package leader and task leader will have to upload their technical reports, test reports, milestones, and deliverables so that they can be accessible by any consortium member quickly. This data could be used by other members for their developments inherent to the project but is protected by the IP, right of use, and confidentiality policy (the “IP Policy”) set up by the consortium in case of any request for external use. The software will be shared through private and secured Gitlab repositories. All personal data will be subject to a written agreement before publication. Along the same lines, we will make sure it complies with the European regulation 2016/679 (GDPR).

### 9.9 Discontinuation Criteria

- Participants may be discontinued from the study under the following circumstances:
- Adverse Events: If a participant experiences severe adverse events or side effects related to the intervention.
- Non-Compliance: If a participant fails to adhere to the study protocol or withdraws consent.
- Other Medical Reasons: If a participant develops a medical condition that contraindicates their continued participation in the study.

### 9.10 Insurance

In accordance with European regulations and ethical guidelines, the study will be insured to cover any potential damages resulting from the research and caused to persons participating in this trial. The insurance coverage aims to ensure the protection and well-being of the trial participants throughout the study. As the sponsor of the clinical trial, MyelinZ is fully committed to adhering to all necessary regulatory requirements and has taken proactive steps to initiate the process of obtaining the required insurance coverage. We will also ensure that all trial participants are protected and provided with the necessary insurance coverage. This ensures they have access to appropriate compensation and support in case of any unforeseen adverse events or damages arising from their participation in the trial.

## 10. Statistical Analysis

### A. Power Analysis Update Sample Size Calculation **Based on:**

- Primary endpoint: EDSS change
- Expected effect size: 0.4
- Power: 90%
- Significance level: 5%
- Dropout rate: 20%

#### Sample Size Justification

**N = 2(Zα/2 + Zβ)²σ²/δ²**

Where:

- α = 0.05 (two-sided)

- β = 0.10 (power = 90%)

- σ = 1.2 (SD of EDSS change)

- δ = 0.4 (minimal clinically important difference)

- Primary MS comparison: n=300

* Effect size: 0.4

* Power: 90%

* Alpha: 0.05

- Healthy Controls: n=100

* Allows detection of differences ≥0.5 SD

* Power: 85%

* Alpha: 0.05 (adjusted for multiple comparisons)

### B. Analysis Plan

- **Sample Size Justification:** Calculated based on EDSS change with a 90% power and significance of 0.05. **Analysis Methods:** Mixed-effects model for repeated measures, intent-to-treat analysis, multiple imputation for missing data.
- Between-Group Comparisons: # Mixed effects model model <- lmer(performance ∼ group * time + age + gender + (1|subject) + (1|site), data = study_data)
- Normative Data Development: # Establish reference ranges norm_ranges <- function(data) { quantiles <-quantile(data, probs = c(0.025, 0.975)) mean_sd <- list(mean = mean(data), sd = sd(data)) return(list(quantiles = quantiles, mean_sd = mean_sd)) }

**Primary Analysis**

- Mixed-effects model for repeated measures
- Intent-to-treat population
- Multiple imputation for missing data

**Secondary Analyses**

1. MSFC score changes
2. MRI volume changes
3. Quality of life measures
4. Adherence patterns
5. Safety outcomes

**Interim Analysis**

- Planned at 12 months
- Safety monitoring
- Futility assessment

**Participant Distribution by Location and Type**

**Figure 1.**
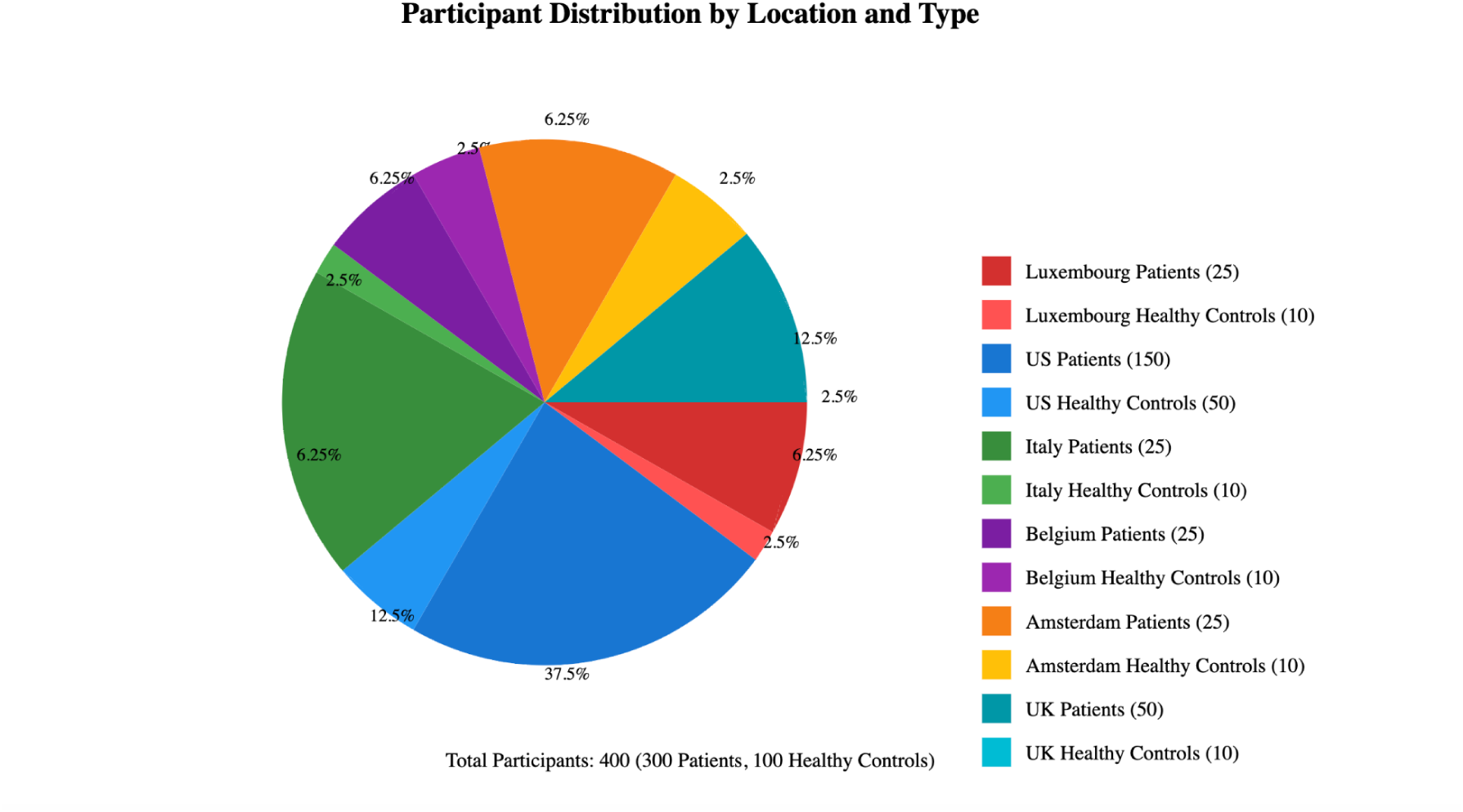
Pie chart. Patient Distribution.

## 11. Ethical Considerations

### Regulatory Compliance

1. ICH-GCP guidelines: Including informed consent and ethics committee approval.
2. Local regulations
3. Ethics committee approval
4. Informed consent

### Patient Protection

1. Privacy measures
2. Confidentiality
3. Withdrawal procedures
4. Patient rights: We will ensure confidentiality, right to withdraw, and patient rights.

## 12. Administrative Aspects

- **Project Timeline:** Major milestones and deadlines.
- **Documentation:** Maintenance of essential documents, regulatory files, and study reports.

## 12. Administrative Aspects

### 12.1 Study Management

1. Steering committee
2. Data monitoring committee
3. Clinical operations
4. Project timeline

### 12.2 Documentation

1. Essential documents
2. Regulatory files
3. Study reports
4. Publications

## 13. Data Privacy

We use Federated Learning (FL), which is a technique that provides privacy protection while enabling the decentralized training of ML models without sharing the local data on nodes but allowing the model parameters coming from all nodes to aggregate a common and more accurate model. However, the model parameters can also be representative of the local data, and they can expose private information about the individuals **[48]** and/or the training data set **[49]**. Therefore, FL is combined with PETs to achieve strong privacy guarantees.

We will also design and implement privacy enhancing technologies to improve security and privacy: The requirements due to compliance with regulations and standards are important drivers for the development and implementation of privacy-enhancing technologies. Particularly, the processing of health data should strictly follow regulations such as the GDPR and HIPAA). To comply with those regulations and many others (i.e., ePrivacy directive), privacy-preserving machine learning solutions have been successfully adopted by researchers, technology developers and innovators in recent years. The idea is, therefore, to integrate machine learning solutions with privacy-enhancing techniques (PETs). However, this integration is not straightforward, and due to the very nature of PETs, the integrated solution may gain in terms of privacy but may lose in terms of ML model performance (utility). Hence, the resulting solution should balance the privacy-utility tradeoff. In the R-MMS project, one of our primary objectives is to identify privacy and security in the application scenario. Then, the PETs module will be designed, developed and integrated with the SaMD solution.

## Measures that are taken to ensure data protection, data ethics, fair data, and open access to the publications

**Data Sharing:** Data and software will be shared through private and secured Gitlab repositories.

**Data Availability:** Data can be made available upon request by the corresponding author in all the planned publications related to this clinical trial and collaborative research project.

**Publications:** We are planning to publish all the results of this work in open-access journals. **Informed Consent Statement:** Informed consent will be obtained from all subjects involved in the study.

**Data Privacy and GDPR Compliance:** MyelinZ has developed an advanced machine learning solution (based on federated learning **[72]**,) to preserve privacy en route to complying with GDPR. This solution first encodes medical records data (based on convolution neural network algorithms), allowing a new mathematical representation of original data. Thereafter, the recorded data will automatically be sent to the hospital’s server, thereby the doctor (and not to the company’s servers) allowing for in-house local training and processing using federated learning. Hence, the company does not directly access or store any patient’s data on its servers, thereby preserving the privacy of patients’ medical data and complying with the general GDPR. Additionally, informed consent explaining the data flow and all the processing will be used during clinical trials and within the mobile app when the product is launched on the market. The concept is summarised in Figure 2.

**Figure 2.**
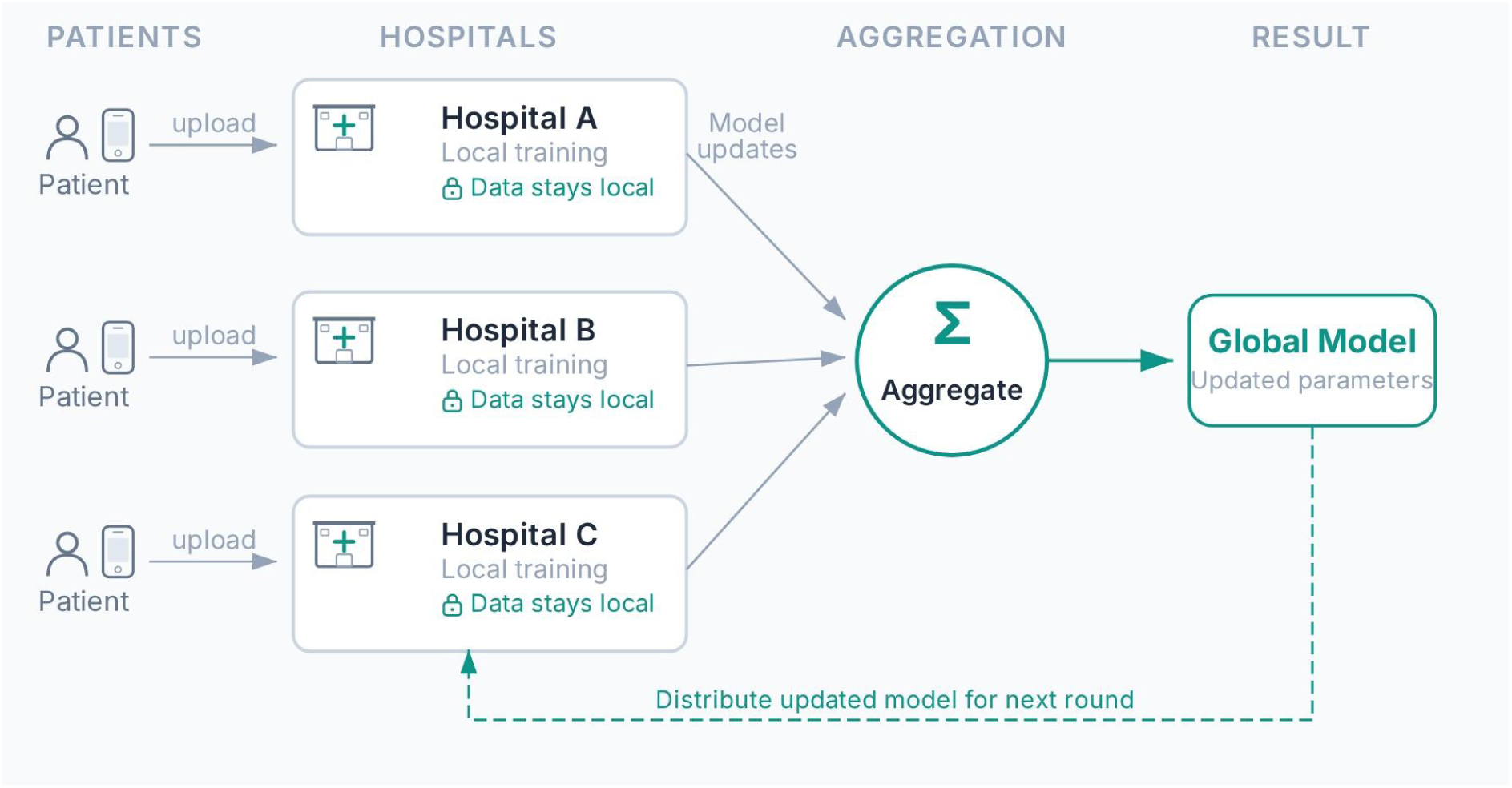
Overview of MyelinZ Federated Learning Concept.

## 14. Benefits and Impact of this Collaborative Clinical & Research Project

**Clinical Impact:** Using this proposed technology, patients and their doctors and therapists can jointly monitor their health status from home, contribute to slowing disease progression, and receive tailored (under-demand) medication depending on their needs. From a health economics standpoint, such software technology will contribute to reducing treatment costs. Moreover, the same SaMD will enable large pharma companies to conduct decentralised & digital clinical trials and develop better-targeted therapies for MS. Large pharma companies are already interested in this SaMD.

**Societal Impact**:

- The proposed clinical project can have several significant societal impacts. Here are a few potential outcomes:
- Improved Access to Care: Remote monitoring can enhance access to healthcare for individuals with MS, particularly those who live in remote areas or have limited mobility. Patients can receive timely care regardless of their geographical location by eliminating the need for frequent in-person visits, improving health outcomes.
- Enhanced Patient Empowerment: Remote monitoring empowers patients by involving them in their care management. By tracking their symptoms, treatment effectiveness, and disease progression remotely, individuals with MS can participate in decision-making, better understand their condition, and make informed choices about their treatment plans.
- Timely Intervention and Personalized Treatments: Remote monitoring can enable healthcare providers to detect disease progression or treatment failures at an early stage. With real-time data and regular updates, medical professionals can intervene promptly, adjust treatment plans, and provide personalized care to patients, optimizing their quality of life and potentially slowing disease progression.
- Increased Efficiency and Cost Savings: Remote monitoring can help healthcare systems achieve greater efficiency and cost savings. By reducing the need for in-person appointments, the burden on healthcare facilities can be alleviated, resulting in shorter patient wait times and potential cost savings for patients and healthcare providers.
- Data-Driven Research and Development: Remote monitoring generates vast amounts of data on MS progression and treatment effectiveness. Aggregated and anonymized data from multiple patients can be used for research and development. This can lead to a deeper understanding of the disease, identifying patterns, and developing new therapies or interventions.
- Remote Clinical Trials and Expanded Research: Remote monitoring can facilitate remote participation in clinical trials for MS treatments. This opens up opportunities for a more diverse and representative participant pool, enabling researchers to gather data from a broader range of individuals with varying demographics and geographical locations. It can also enhance participant retention and reduce the burden associated with travel and in-person visits.
- Improved Quality of Life: By enabling remote monitoring, individuals with MS can experience enhanced overall quality of life. They can reduce the stress and inconvenience of frequent clinic visits, maintain independence, and engage in daily activities with minimal disruption. This can positively impact their emotional well-being, productivity, and social interactions.

## 15. Appendix

### BodyMirror Sensor

BodyMirror sensor is a compact, wearable EEG headband with four dry electrodes, designed to record brain activity at a 250 Hz sampling frequency. It features an integrated electronic module that amplifies, digitises, and transmits the EEG signals to the cloud using Bluetooth LE, allowing for secure data storage and access.

The BodyMirror has the following electrodes:

- O1, O2, T3, T4 (names according to the 10-20 international scheme of placement of electrodes) recording electrodes.
- FpZ – reference electrode.
- GND – common electrode.

The T3 and T4 electrodes provide direct contact with the scalp in the temporal lobe areas, and the O1 and O2 electrodes in the occipital region. The signals are recorded in monopolar method (O1 – FpZ, O2 – FpZ, T3 – FpZ, T4 – FpZ).

### Sensor connection and positioning

Here you can find the step-by-step instructions on how to turn on, connect, and put on the EEG headset:

- Stretch the textile band in the clasp area.
- Place BodyMirror on your head, considering: 1) the location of the reference and general electrodes on the forehead; 2) the direction of the arrows in the area of the common electrode (should be directed upwards); 3)the location of the T3 and T4 electrodes in the temporal region, and the O1 and O2 electrodes in the occipital region.
- Tighten the textile band according to the size of the head.

### MuscleMirror Sensor

MuscleMirror is a wearable 1-channel electromyography (EMG) and IMU sensor. The sensor uses 1 EMG dry electrode and 6-axis motion sensor to collect real-time EMG and IMU raw data respectively. The EMG data is recorded at 1000Hz, instead the data from the IMU sensor at 95Hz.

The characteristics of the power supply are:

- Input parameters: 100–240 VAC, 50/60 Hz, 0.1–0.5 A (or in accordance with your region).
- Output parameters: 5V, DC (direct current) 1–2 A.

### Sensor connection and positioning

Here you can find the step-by-step instructions on how to turn on, connect, and put on the MuscleMirror:

- Take the sensor and put it on your forearm. Make sure the part of the sensor with the power button is placed directly on your muscle.
- Press the power button to turn the sensor on. The sensor is turned on when the green light is flickering.
- Make sure your arm is not leaned against anything (table, leg, etc.).

### Data Security and Privacy Measures

- **Data Security and Privacy Compliance**: The protocol complies with all GDPR and HIPAA regulations for data protection. The BodyMirror system utilises federated learning to maintain data privacy, enabling decentralised data processing on local devices. Encryption protocols are applied to all data transfers, and additional Privacy-Enhancing Technologies (PETs) have been integrated. A Data Protection Officer (DPO) will oversee compliance throughout the trial, ensuring regular audits and privacy assessments. A secure electronic case report form (eCRF) platform will log and backup all data, accessible only to authorised personnel with unique credentials.

### Patient Consent Forms and Information Sheets

- **Patient Information Sheet**: This document will inform participants about the study’s purpose, the use of wearable devices, the data collection process, and the specific health and personal information that will be recorded. Participants will be informed of their rights, including the right to withdraw at any point without consequences.
- **Consent Form Language**: Consent forms will include specific agreements from participants regarding data sharing for research purposes, the use of anonymized data in publications, and compliance with privacy regulations (GDPR/HIPAA). Additionally, it will detail any possible discomforts or risks related to wearing devices, data privacy, and procedures for discontinuing participation.

## 3. Device and Software Validation

Prior feasibility testing involving 15 participants demonstrated a **>95% successful data capture rate** and high user adherence. Device signal quality was benchmarked against clinical reference standards to confirm the reliability of EEG and EMG recordings in a home-based environment. These findings supported progression to the current randomised clinical trial.

- **Device Validation Statement**: The EEG, EMG, and IMU devices in the BodyMirror system have undergone rigorous testing to ensure accuracy, usability, and safety. Validation studies have shown [brief summary of validation metrics, e.g., ’EEG recordings demonstrate accuracy within ±2% of clinical standards’]. All devices are compliant with ISO 13485 medical device quality standards.
- **Software Testing Summary**: The MyelinFace app and MyelinBoard dashboard underwent usability testing with a small cohort of MS patients, confirming ease of use, stability, and data integrity during data capture. Preliminary data demonstrated reliability across different operating systems (iOS, Android). Additionally, machine learning algorithms within MyelinFace meet accuracy standards (specificity and sensitivity measures where applicable).

## 4. Pilot Feasibility Testing

- **Pilot Testing Summary**: A preliminary feasibility study was conducted with 15 participants (MS patients and healthy controls) to validate system operations. Results indicated over 95% data capture success rate with minimal dropout due to technical challenges. Feedback on the game interface confirmed user engagement and ease of use, with adjustments made to optimize game timing and difficulty.
- **Impact on Protocol**: Based on pilot feedback, session timing and device calibration have been optimized, ensuring better patient adherence. Adjustments to consent forms and instructions were also implemented to enhance clarity.

## 5. Protocol for Handling Adverse Events and Discontinuation

- **Adverse Event Management Protocol**: Any adverse events, including device-related discomfort or psychological distress, will be documented and reviewed by an independent monitoring committee. Participants experiencing adverse events will receive immediate support and, if necessary, be withdrawn from the study.
- **Discontinuation Protocol**: Participants can voluntarily withdraw at any time, or be withdrawn by the study team if they experience severe adverse events. All discontinued cases will be logged, with reasons documented and analyzed to assess protocol adherence and safety.

## 6. Training and Standardization for Multi-Site Consistency

- **Training Program Summary**: All clinical sites involved will undergo a comprehensive training program covering device usage, data collection methods, and patient handling. A standardized training manual and online modules will be provided, ensuring consistent protocol adherence across sites.
- **Monitoring and Quality Assurance**: “Regular site visits and audits will verify compliance with trial standards. Standard operating procedures (SOPs) will be implemented, covering everything from patient onboarding to data logging and adverse event reporting. Monthly inter-site meetings will allow feedback and protocol adjustments, fostering consistency.

**List of Abbreviations**
BMRM: BodyMirror Remote Monitoring
EDSS: Expanded Disability Status Scale
MS: Multiple Sclerosis
MRI: Magnetic Resonance Imaging
EMG: Electromyography
EEG: Electroencephalography
IMU: Inertial Measurement Unit
MS: Multiple Sclerosis
MSFC: Multiple Sclerosis Functional Composite
RRMS: Relapsing-Remitting Multiple Sclerosis
T25FW: Timed 25-Foot Walk
9HPT: 9-Hole Peg Test
PASAT: Paced Auditory Serial Addition Test
SaMD: Software-as-a-medical device
RRMS: Relapsing-remitting MS
MEP: Motor evoked potentials
SSEP: Somatosensory evoked potentials
VEP: Visual evoked potential
IMU: Inertial measurement unit
PMS: Progressive MS
DSI: Dysphonia Severity Index
PwMS: Patient with MS
MSFC: Multiple Sclerosis Functional Composite
DMT: Disease Modifying Therapy
SPMS: Secondary Progressive MS
GDPR: General Data Protection Regulation
PETs: Privacy Enhancing Technologies
HE: Homomorphic Encryption
MPC: Secure Multiparty Computation
kFLCs: Kappa-free light chains
MFZ: Measles-rubella-zoster
NLC: Neurofilament light chains
HIPAA: Health Insurance Portability and Accountability Act
RPM: Remote Patient Monitoring
PETs: Privacy-Enhancing Techniques

## Data Availability

All data produced in the present study are available upon reasonable request to the authors

## Notes

### Competing Interest Statement

The authors have declared no competing interest.

### Funding Statement

This work was supported in part by a PhD grant from the Luxembourg National Research Fund (FNR) under the project reference 17223919/MMS/Industrial Fellowship.

## References

1. Walton, C. et al. Rising prevalence of multiple sclerosis worldwide: Insights from the atlas of ms, third edition. Multiple Scler. J. 26, 1816–1821 (2020).

2. Filippi M, Bar-Or A, Piehl F, Preziosa P, Solari A, Vukusic S, Rocca MA. Multiple sclerosis. Nat Rev Dis Primers. 2018 Nov 8;4(1):43. doi: 10.1038/s41572-018-0041-4. Erratum in: Nat Rev Dis Primers. 2018 Nov 22;4(1):49. PMID: 30410033.

3. https://multiplesclerosisnewstoday.com/news-posts/2023/01/17/early-progression-independent-relapses-linked-worse-disability/ Online access [04.07.2023].

4. Engelhard MM, S. K. L. J. G. M., Patek SD. Remotely engaged: Lessons from remote monitoring in multiple sclerosis. Int. journal medical informatics 100 (2017).

5. https://www.nationalmssociety.org/Treating-MS/Medications Online access [15.06.2023].

6. Cagol A, Schaedelin S, Barakovic M, et al. Association of Brain Atrophy With Disease Progression Independent of Relapse Activity in Patients With Relapsing Multiple Sclerosis. JAMA Neurol. 2022;79(7):682–692. doi:10.1001/jamaneurol.2022.1025

7. Kathiravan S, Kanakaraj J. A review on potential issues and challenges in MR imaging. ScientificWorldJournal. 2013 Nov 27;2013:783715. doi: 10.1155/2013/783715. PMID: 24381523; PMCID: PMC3863452.

8. Weinstock-Guttman B, Sormani MP, Repovic P. Predicting Long-term Disability in Multiple Sclerosis: A Narrative Review of Current Evidence and Future Directions. Int J MS Care. 2022 Jul-Aug;24(4):184–188. doi: 10.7224/1537-2073.2020-114. Epub 2022 Oct 5. PMID: 35875463; PMCID: PMC9296054.

9. Hauser SL, Cree BAC. Treatment of Multiple Sclerosis: A Review. Am J Med. 2020 Dec;133(12):1380–1390.e2. doi: 10.1016/j.amjmed.2020.05.049. Epub 2020 Jul 17. PMID: 32682869; PMCID: PMC7704606.

10. https://www.healthline.com/health/multiple-sclerosis/drugs#talk-with-a-doctor Online access [01.06.2023].

11. Gafson A, Craner MJ, Matthews PM. Personalised medicine for multiple sclerosis care. Mult Scler. 2017 Mar;23(3):362–369. doi: 10.1177/1352458516672017. Epub 2016 Sep 28. PMID: 27672137.

12. Pathak, L. (2023). ’Personalized Treatment for Multiple Sclerosis: The Role of Precision Medicine’, Neurology Letters, 2(1), pp. 30–34. doi: 10.52547/nl.2.1.30

13. Chitnis T, Prat A. A roadmap to precision medicine for multiple sclerosis. Mult Scler. 2020 Apr;26(5):522–532. doi: 10.1177/1352458519881558. Epub 2020 Jan 22. PMID: 31965904.

14. Saleem S, Anwar A, Fayyaz M, Anwer F, Anwar F. An Overview of Therapeutic Options in Relapsing-remitting Multiple Sclerosis. Cureus. 2019 Jul 26;11(7):e5246. doi: 10.7759/cureus.5246. PMID: 31565644; PMCID: PMC6759037.

15. Engel, S., Zipp, F. Preventing disease progression in multiple sclerosis—insights from large real-world cohorts. Genome Med 14, 41 (2022). 10.1186/s13073-022-01044-8

16. The Economic Burden of Multiple Sclerosis in the United States Estimate of Direct and Indirect Costs Bruce Bebo, Inna Cintina, Nicholas LaRocca, Leslie Ritter, Bari Talente, Daniel Hartung, Surachat Ngorsuraches, Mitchell Wallin, Grace Yang Neurology May 2022, 98 (18) e1810–e1817; DOI: 10.1212/WNL.0000000000200150

17. https://www.ajmc.com/view/cost-effectiveness-multiple-sclerosis-economic-burden Online access [01.07.2023].

18. van der Walt A, Butzkueven H, Shin RK, Midaglia L, Capezzuto L, Lindemann M, Davies G, Butler LM, Costantino C, Montalban X. Developing a Digital Solution for Remote Assessment in Multiple Sclerosis: From Concept to Software as a Medical Device. Brain Sci. 2021 Sep 21;11(9):1247. doi: 10.3390/brainsci11091247. PMID: 34573267; PMCID: PMC8471038.

19. Leocani, L. et al. Electroencephalographic coherence analysis in multiple sclerosis: correlation with clinical, neuropsychological, and MRI findings. 69, 192–198 (2000)

20. H., P. J. Electromyographic findings in multiple sclerosis: remitting signs of denervation. Muscle nerve 5, 157–160 (1982).

21. Coca-Tapia M, Cuesta-Gómez A, Molina-Rueda F, Carratalá-Tejada M. Gait Pattern in People with Multiple Sclerosis: A Systematic Review. Diagnostics (Basel). 2021 Mar 24;11(4):584. doi: 10.3390/diagnostics11040584. PMID: 33805095; PMCID: PMC8064080.

22. Behrenbeck, Jan et al. “Classification and regression of spatio-temporal signals using NeuCube and its realization on SpiNNaker neuromorphic hardware.” Journal of Neural Engineering 16 (2019)

23. Z. Tayeb, E. Erçelik and J. Conradt, “Decoding of motor imagery movements from EEG signals using SpiNNaker neuromorphic hardware,” 2017 8th International IEEE/EMBS Conference on Neural Engineering (NER), Shanghai, China, 2017, pp. 263-266, doi: 10.1109/NER.2017.8008341.

24. Tayeb Z, Waniek N, Fedjaev J, Ghaboosi N, Rychly L, Widderich C, Richter C, Braun J, Saveriano M, Cheng G, Conradt J. Gumpy: a Python toolbox suitable for hybrid brain-computer interfaces. J Neural Eng. 2018 Dec;15(6):065003. doi: 10.1088/1741-2552/aae186. Epub 2018 Sep 14. PMID: 30215610.

25. Tayeb, Z., Bose, R., Dragomir, A. et al. Decoding of Pain Perception using EEG Signals for a Real-Time Reflex System in Prostheses: A Case Study. Sci Rep 10, 5606 (2020). 10.1038/s41598-020-62525-7

26. Picton TW. The P300 wave of the human event-related potential. J Clin Neurophysiol. 1992 Oct;9(4):456–79. doi: 10.1097/00004691-199210000-00002. PMID: 1464675.

27. Klistorner A, Graham SL. Role of Multifocal Visually Evoked Potential as a Biomarker of Demyelination, Spontaneous Remyelination, and Myelin Repair in Multiple Sclerosis. Front Neurosci. 2021 Oct 29;15:725187. doi: 10.3389/fnins.2021.725187. PMID: 34776840; PMCID: PMC8586643.

28. Kim H, Gao S, Yi B, Shi R, Wan Q, Huang Z. Validation of the Dysphonia Severity Index in the Dr. Speech Program. J Voice. 2019 Nov;33(6):948.e23-948.e29. doi: 10.1016/j.jvoice.2019.08.011. Epub 2019 Sep 13. PMID: 31526665.

29. Hossen A, Anwar AR, Koirala N, Ding H, Budker D, Wickenbrock A, Heute U, Deuschl G, Groppa S, Muthuraman M. Machine learning aided classification of tremor in multiple sclerosis. EBioMedicine. 2022 Aug;82:104152. doi: 10.1016/j.ebiom.2022.104152. Epub 2022 Jul 11. PMID: 35834887; PMCID: PMC9287478.

30. Mele, G. et al. Simultaneous EEG-fMRI for functional neurological assessment. Front. Neurol. 10 (2019).

31. Capone F, Motolese F, Falato E, Rossi M, Di Lazzaro V. The Potential Role of Neurophysiology in the Management of Multiple Sclerosis-Related Fatigue. Front Neurol. 2020 Apr 22;11:251. doi: 10.3389/fneur.2020.00251. PMID: 32425869; PMCID: PMC7212459.

32. University of California, San Francisco MS-EPIC Team; Cree BAC, Hollenbach JA, Bove R, Kirkish G, Sacco S, Caverzasi E, Bischof A, Gundel T, Zhu AH, Papinutto N, Stern WA, Bevan C, Romeo A, Goodin DS, Gelfand JM, Graves J, Green AJ, Wilson MR, Zamvil SS, Zhao C, Gomez R, Ragan NR, Rush GQ, Barba P, Santaniello A, Baranzini SE, Oksenberg JR, Henry RG, Hauser SL. Silent progression in disease activity-free relapsing multiple sclerosis. Ann Neurol. 2019 May;85(5):653–666. doi: 10.1002/ana.25463. Epub 2019 Mar

30. PMID: 30851128; PMCID: PMC6518998.

33. Chilińska A, Ejma M, Turno-Kręcicka A, Guranski K, Misiuk-Hojlo M. Analysis of retinal nerve fibre layer, visual evoked potentials and relative afferent pupillary defect in multiple sclerosis patients. Clin Neurophysiol. 2016 Jan;127(1):821–826. doi: 10.1016/j.clinph.2015.06.025. Epub 2015 Jul 4. PMID: 26251105.

34. Green AJ, Gelfand JM, Cree BA, Bevan C, Boscardin WJ, Mei F, Inman J, Arnow S, Devereux M, Abounasr A, Nobuta H, Zhu A, Friessen M, Gerona R, von Büdingen HC, Henry RG, Hauser SL, Chan JR. Clemastine fumarate as a remyelinating therapy for multiple sclerosis (ReBUILD): a randomised, controlled, double-blind, crossover trial. Lancet. 2017 Dec 2;390(10111):2481–2489. doi: 10.1016/S0140-6736(17)32346-2. Epub 2017 Oct 10. PMID: 29029896.

35. Genç G, Demirkaya Ş, Bek S, Odabaşi Z. Clinical, Radiological and Electrophysiological Comparison of Immunomodulatory Therapies in Multiple Sclerosis. Noro Psikiyatr Ars. 2017 Jun;54(2):116–124. doi: 10.5152/npa.2016.12621. Epub 2016 Mar 1. PMID: 28680308; PMCID: PMC5491660.

36. Kiiski HS, Ní Riada S, Lalor EC, Gonçalves NR, Nolan H, Whelan R, Lonergan R, Kelly S, O’Brien MC, Kinsella K, Bramham J, Burke T, Ó Donnchadha S, Hutchinson M, Tubridy N, Reilly RB. Delayed P100-Like Latencies in Multiple Sclerosis: A Preliminary Investigation Using Visual Evoked Spread Spectrum Analysis. PLoS One. 2016 Jan 4;11(1):e0146084. doi: 10.1371/journal.pone.0146084. PMID: 26726800; PMCID: PMC4699709.

37. Calugaru L, Calugaru GT, Calugaru OM. Evoked Potentials in Multiple Sclerosis Diagnosis and Management. Curr Health Sci J. 2016 Oct-Dec;42(4):385–389. doi: 10.12865/CHSJ.42.04.08. Epub 2016 Feb 28. PMID: 30581593; PMCID: PMC6269618.

38. Covey TJ, Golan D, Doniger GM, Sergott R, Zarif M, Srinivasan J, Bumstead B, Wilken J, Buhse M, Mebrahtu S, Gudesblatt M. Visual evoked potential latency predicts cognitive function in people with multiple sclerosis. J Neurol. 2021 Nov;268(11):4311–4320. doi: 10.1007/s00415-021-10561-2. Epub 2021 Apr 18. PMID: 33870445.

39. Hameau S, Bensmail D, Roche N, Zory R. Fatigability in Patients With Multiple Sclerosis During Maximal Concentric Contractions. Arch Phys Med Rehabil. 2017 Jul;98(7):1339–1347. doi: 10.1016/j.apmr.2016.12.014. Epub 2017 Jan 25. PMID: 28130080.

40. Krysko KM, Akhbardeh A, Arjona J, Nourbakhsh B, Waubant E, Antoine Gourraud P, Graves JS. Biosensor vital sign detects multiple sclerosis progression. Ann Clin Transl Neurol. 2021 Jan;8(1):4–14. doi: 10.1002/acn3.51187. Epub 2020 Nov 19. PMID: 33211403; PMCID: PMC7818086.

41. Kanzler CM, Sylvester R, Gassert R, Kool J, Lambercy O, Gonzenbach R. Goal-directed upper limb movement patterns and hand grip forces in multiple sclerosis. Mult Scler J Exp Transl Clin. 2022 Aug 11;8(3):20552173221116272. doi: 10.1177/20552173221116272. PMID: 35982915; PMCID: PMC9380226.

42. Huang SC, Guerrieri S, Dalla Costa G, Pisa M, Leccabue G, Gregoris L, Comi G, Leocani L. Intensive Neurorehabilitation and Gait Improvement in Progressive Multiple Sclerosis: Clinical, Kinematic and Electromyographic Analysis. Brain Sci. 2022 Feb 12;12(2):258. doi: 10.3390/brainsci12020258. PMID: 35204021; PMCID: PMC8870152.

43. Svoboda E, Bořil T, Rusz J, Tykalová T, Horáková D, Guttmann CRG, Blagoev KB, Hatabu H, Valtchinov VI. Assessing clinical utility of machine learning and artificial intelligence approaches to analyze speech recordings in multiple sclerosis: A pilot study. Comput Biol Med. 2022 Sep;148:105853. doi: 10.1016/j.compbiomed.2022.105853. Epub 2022 Jul 15. PMID: 35870318.

44. O’Keeffe C, Yap SM, Davenport L, Cogley C, Craddock F, Kennedy A, Tubridy N, Looze C, Suleyman N, O’Keeffe F, Reilly RB, McGuigan C. Association between speech rate measures and cognitive function in people with relapsing and progressive multiple sclerosis. Mult Scler J Exp Transl Clin. 2022 Aug 17;8(3):20552173221119813. doi: 10.1177/20552173221119813. PMID: 36003923; PMCID: PMC9393591.

45. Rusz J, Vaneckova M, Benova B, Tykalova T, Novotny M, Ruzickova H, Uher T, Andelova M, Novotna K, Friedova L, Motyl J, Kucerova K, Krasensky J, Horakova D. Brain volumetric correlates of dysarthria in multiple sclerosis. Brain Lang. 2019 Jul;194:58–64. doi: 10.1016/j.bandl.2019.04.009. Epub 2019 May 15. PMID: 31102976.

46. Fazeli M, Moradi N, Soltani M, Naderifar E, Majdinasab N, Latifi SM, Dastoorpour M. Dysphonia Characteristics and Vowel Impairment in Relation to Neurological Status in Patients with Multiple Sclerosis. J Voice. 2020 May;34(3):364–370. doi: 10.1016/j.jvoice.2018.09.018. Epub 2018 Oct 19. PMID: 30342799.

47. Nordio S, Bernitsas E, Meneghello F, Palmer K, Stabile MR, Dipietro L, Di Stadio A. Expiratory and phonation times as measures of disease severity in patients with Multiple Sclerosis. A case-control study. Mult Scler Relat Disord. 2018 Jul;23:27–32. doi: 10.1016/j.msard.2018.04.010. Epub 2018 Apr 21. PMID: 29753215.

48. Fredrikson, Matt, Somesh Jha and Thomas Ristenpart. “Model Inversion Attacks that Exploit Confidence Information and Basic Countermeasures.” Proceedings of the 22nd ACM SIGSAC Conference on Computer and Communications Security (2015):

49. R. Shokri, M. Stronati, C. Song and V. Shmatikov, “Membership Inference Attacks Against Machine Learning Models,” 2017 IEEE Symposium on Security and Privacy (SP), San Jose, CA, USA, 2017, pp. 3–18, doi: 10.1109/SP.2017.41.

50. Craig Gentry. 2009. Fully homomorphic encryption using ideal lattices. In Proceedings of the forty-first annual ACM symposium on Theory of computing (STOC ’09). Association for Computing Machinery,, 169–178. 10.1145/1536414.1536440

51. A. C. -C. Yao, “How to generate and exchange secrets,” 27th Annual Symposium on Foundations of Computer Science (sfcs 1986), Toronto, ON, Canada, 1986, pp. 162–167, doi: 10.1109/SFCS.1986.25.

52. Samarati, Pierangela and Latanya Sweeney. “Protecting privacy when disclosing information: k-anonymity and its enforcement through generalization and suppression.” (1998).

53. Martin Abadi, Andy Chu, Ian Goodfellow, H. Brendan McMahan, Ilya Mironov, Kunal Talwar, and Li Zhang. 2016. Deep Learning with Differential Privacy. In Proceedings of the 2016 ACM SIGSAC Conference on Computer and Communications Security (CCS ’16). Association for Computing Machinery, New York, NY, USA, 308–318. 10.1145/2976749.2978318

54. A. Machanavajjhala, J. Gehrke, D. Kifer and M. Venkitasubramaniam, “L-diversity: privacy beyond k-anonymity,” 22nd International Conference on Data Engineering (ICDE’06), Atlanta, GA, USA, 2006, pp. 24–24, doi: 10.1109/ICDE.2006.1.

55. N. Li, T. Li and S. Venkatasubramanian, “t-Closeness: Privacy Beyond k-Anonymity and l-Diversity,” 2007 IEEE 23rd International Conference on Data Engineering, Istanbul, Turkey, 2007, pp. 106–115, doi: 10.1109/ICDE.2007.367856.

56. Cynthia Dwork and Aaron Roth. 2014. The Algorithmic Foundations of Differential Privacy. Found. Trends Theor. Comput. Sci. 9, 3–4 (August 2014), 211–407. 10.1561/0400000042

57. K. Wei et al., “Federated Learning With Differential Privacy: Algorithms and Performance Analysis,” in IEEE Transactions on Information Forensics and Security, vol. 15, pp. 3454–3469, 2020, doi: 10.1109/TIFS.2020.2988575.

58. Hao Chen, Wei Dai, Miran Kim, and Yongsoo Song. 2019. Efficient Multi-Key Homomorphic Encryption with Packed Ciphertexts with Application to Oblivious Neural Network Inference. In Proceedings of the 2019 ACM SIGSAC Conference on Computer and Communications Security (CCS ’19). Association for Computing Machinery, New York, NY, USA, 395–412. 10.1145/3319535.3363207

59. Zhang, Q., Jing, S., Zhao, C., Zhang, B., Chen, Z. (2022). Efficient Federated Learning Framework Based on Multi-Key Homomorphic Encryption. In: Barolli, L. (eds) Advances on P2P, Parallel, Grid, Cloud and Internet Computing. 3PGCIC 2021. Lecture Notes in Networks and Systems, vol 343. Springer, Cham. 10.1007/978-3-030-89899-1_10

60. R. Kanagavelu, et al., “Two-Phase Multi-Party Computation Enabled Privacy-Preserving Federated Learning,” in 2020 20th IEEE/ACM International Symposium on Cluster, Cloud and Internet Computing (CCGRID), Melbourne, Australia, 2020 pp. 410–419.

61. Vázquez-Marrufo, M. et al. Quantitative electroencephalography reveals different physiological profiles between benign and remitting-relapsing multiple sclerosis patients. BMC neurology 8, 44 (2008).

62. Torabi, A., Daliri, M. R. & Sabzposhan, S. Diagnosis of multiple sclerosis from EEG signals using nonlinear methods. Australas. physical engineering sciences medicine 40 (2017).

63. Leocani, L. et al. Electroencephalographic coherence analysis in multiple sclerosis: correlation with clinical, neuropsychological, and MRI findings. 69, 192–198 (2000).

64. Luzzio, C. & Dangond, F. https://www.medscape.com/answers/1146199-5787/what-is-the-role-of-eeg-in-the-evaluationof-multiple-sclerosis-ms. Online access: 15-04-2022.

65. Kiiski, H. et al. Only low frequency event-related EEG activity is compromised in multiple sclerosis: Insights from an independent component clustering analysis. PloS one 7, e45536 (2012).

66. Karaca, B. K., Aksahin, M. F. & Öcal, R. Detection of multiple sclerosis disease by EEG coherence analysis. In 2019. Medical Technologies Congress (TIPTEKNO), 1–4 (2019).

67. Leocani, L. et al. EEG correlates of cognitive impairment in ms. The Italian J. Neurol. Sci. 19, S413–S417 (2004).

69. Kim, Youngeun, Yeshwanth Venkatesha and Priyadarshini Panda. “PrivateSNN: Privacy-Preserving Spiking Neural Networks.” AAAI Conference on Artificial Intelligence (2021).

69. Ali Rasteh, Florian Delpech, Carlos Aguilar-Melchor, Romain Zimmer, Saeed Bagheri Shouraki, Timothée Masquelier, Encrypted internet traffic classification using a supervised spiking neural network, Neurocomputing, Volume 503, 2022, Pages 272-282,

70. Jihang Wang, Dongcheng Zhao, Guobin Shen, Qian Zhang, Yi Zeng, “DPSNN: A Differentially Private Spiking Neural Network with Temporal Enhanced Pooling”, arXiv:2205.12718

71. Y. Venkatesha, Y. Kim, L. Tassiulas and P. Panda, “Federated Learning With Spiking Neural Networks,” in IEEE Transactions on Signal Processing, vol. 69, pp. 6183–6194, 2021, doi: 10.1109/TSP.2021.3121632.

72. Abreha HG, Hayajneh M, Serhani MA. Federated Learning in Edge Computing: A Systematic Survey. Sensors (Basel). 2022 Jan 7;22(2):450. doi: 10.3390/s22020450. PMID: 35062410; PMCID: PMC8780479.

